# Towards an open analysis ecosystem for *Plasmodium* genomic epidemiology

**DOI:** 10.1101/2025.04.01.25325032

**Authors:** Shazia Ruybal-Pesántez, Jorge Amaya-Romero, Sophie Bérubé, Nicholas F. Brazeau, Mouhamadou Fadel Diop, Nicholas Hathaway, Jason Hendry, Kirsty McCann, Kathryn Murie, Maxwell Murphy, Karamoko Niaré, Jody Phelan, Stephen F. Schaffner, Alfred Simkin, Aimee R. Taylor, Bryan Greenhouse, Amy Wesolowski, Robert Verity

## Abstract

Major advances in *Plasmodium* sequencing approaches, bioinformatic pipelines, and data analysis tools have provided valuable insights into malaria epidemiology from parasite genomic data. However, translating genetic data into actionable information for decision-makers remains a challenge. Significant barriers limit the integration of these advances into a functional data analysis ecosystem that produces standardized, interpretable results for use by national malaria control programs. The *Plasmodium* Genomic Epidemiology (PlasmoGenEpi) network convened 18 subject matter experts across 15 institutions at the **R**eproducibility, **A**ccessibility, **D**ocumentation, and **I**nteroperability **S**tandards **H**ackathon in 20**23 (RADISH23)** to identify available analysis tools, evaluate software standards, improve documentation, and outline workflows. Eight use cases for genomic data were identified, and a subset were developed into analysis workflows in terms of a series of connected functionalities. Software tools were then mapped against functionalities to outline a modular approach to data analysis for these use cases. In addition to outlining workflows, a set of objective criteria were developed for evaluating software standards. Forty *Plasmodium* genomic analysis tools were identified, of which 22 were prioritized for software standards evaluation. Additional tutorials were developed for 10 tools in the form of reproducible code applied to shared datasets. These resources are available on **PGEforge** (<u>mrc-</u> <u>ide.github.io/PGEforge</u>), a new community resource that serves as a central, open repository for current and future resources for malaria genomic data analysis.

## Introduction

### The central role of data analysis in malaria genomics

In the two decades since sequencing the first *Plasmodium falciparum* genome (Gardner et al., 2002), the malaria research community has devised a wide array of methods aimed at generating insights into malaria epidemiology and transmission dynamics from parasite genetic data. Laboratories can now sequence whole genomes or obtain deep coverage of many genomic targets from small volumes of blood; advanced bioinformatic pipelines can detect minority alleles present in polyclonal infections while removing errors; an array of computational tools have been developed to distill critical insights from the data; and a variety of visualization methods have been created to communicate results to stakeholders. Collectively, these advancements provide the foundation for establishing a virtuous cycle linking public health personnel with laboratory and data scientists (Figure 1). This collaboration is essential for designing meaningful studies, generating and analyzing sequence data, and translating results into actionable information. An integrated approach will be critical when responding to new threats, including the emergence and spread of drug and diagnostic resistance, increased transmission due to insecticide resistance and invasive mosquito species, and importation due to human migration. However, several significant barriers currently hinder the integration of these efforts into a cohesive data generation and analysis ecosystem.

**Figure 1.**
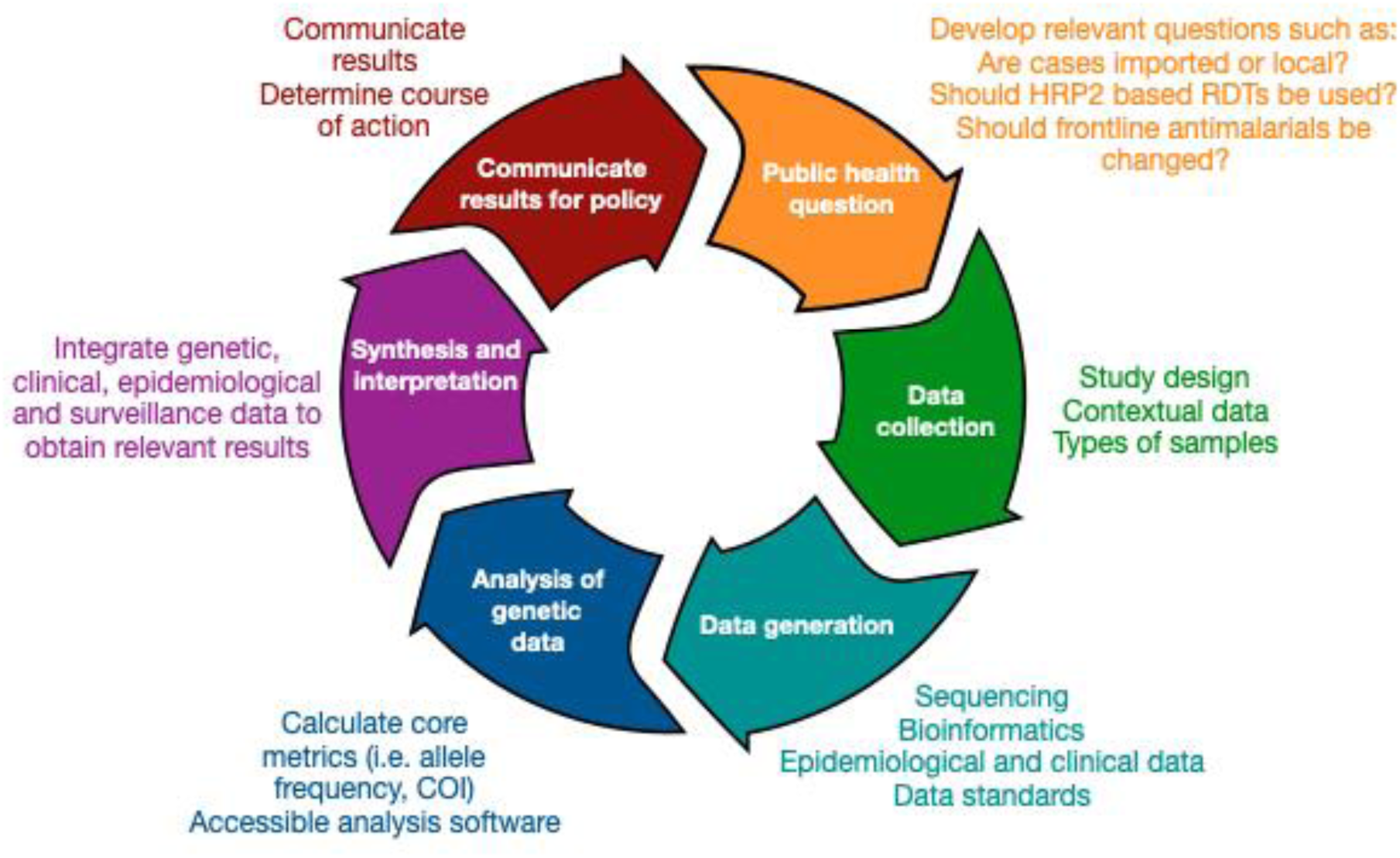
Model for an ideal genomic epidemiology framework involving public health stakeholders and scientists. This framework outlines the iterative cycle of malaria genomic epidemiology, integrating public health priority questions, data collection and generation, genetic data analysis, and synthesis and interpretation to inform malaria control and elimination policy. The process begins with defining public health questions relevant to national malaria control and elimination programmes, such as whether cases are imported or locally acquired, whether HRP2-based rapid diagnostic tests (RDTs) should continue to be used, or whether frontline antimalarial drugs remain effective. Next, data collection involves appropriate study design and sample types, as well as contextual data, for example demographic information or travel history data. Subsequently, data generation will involve sequencing and bioinformatic processing, as well as epidemiological and clinical data, such as light microscopy and PCR diagnosis of malaria infections. Robust data standards would enable harmonization of downstream analysis of genetic data. This stage includes derivation of key metrics from genetic data, such as allele frequencies and complexity of infection (COI), requiring accessible analysis software. The results are then synthesized and interpreted by integrating genetic, clinical, epidemiological and surveillance data to extract relevant insights and determine appropriate courses of action. Findings are then communicated to public-health decision makers and programme officers, guiding intervention strategies and informing further research questions, thereby continuing the cycle.

Substantial variability in sequencing, bioinformatic methods, and nonstandard data formats, create challenges in harmonizing downstream analysis. This variability, however, should not impede progress; rather, adopting a principled approach to analysis should enable users to select from various data generation options while ensuring the comparability of final results. Many analysis tools have been created over the past 15 years to address the challenges specific to analyzing *Plasmodium* genetic data. However, these tools have not been systematically assessed, and vary widely in their adherence to best practices in software development, for example FAIR standards (Barker et al., 2022) or those proposed by the Public Health Alliance for Genomic Epidemiology (PH4GE) (Libuit KG, et al., 2023). Moreover, there are few, if any, standards for data exchange between them, limiting interoperability. A unified ecosystem that prioritizes interoperability and accessibility with clear data-sharing guidelines and modular workflows, as proposed for public health genomic surveillance more broadly (Black et al., 2020), is essential to maximize the impact of malaria genomic data. Rather than investing resources in self-contained analysis pipelines, a more sustainable and effective solution would be to create a collaborative, transparent, and open platform of curated interoperable software to translate genetic and contextual data into meaningful results. The advantages of harmonization are clearly visible in other related fields, such as public health genomic surveillance of antimicrobial resistance, where community-led efforts to evaluate and unify software and data standards have led to wide uptake and a clear path to improve interoperability of workflows (Mendes et al., 2024).

Previous community efforts to define applications of genetic epidemiological methods for answering research questions and informing malaria policy have resulted in seven defined use cases (Dalmat et al., 2019). Implementing workflows for these applications requires going a step further and outlining the types of analyses needed to translate genetic data to desired results. Understanding which tools are available to robustly perform which functions would aid the development of modular analysis workflows.

As a first step in addressing these needs, the Plasmodium Genomic Epidemiology (PlasmoGenEpi) network convened 18 subject matter experts from 14 institutions to catalogue and test available software tools, evaluate software standards and usability, improve documentation, and evaluate next steps in the context of key use cases for malaria public health and research. This took place at the Reproducibility, Accessibility, Documentation, and Implementation Software Hackathon in 2023 (RADISH23) - a four-day event hosted by Johns Hopkins University. This manuscript outlines the main outputs from this event, which center around the PGEforge website – a new community resource for malaria genomic analysis tools.

## Results

### Use cases and analysis workflows for *Plasmodium* genomic epidemiology

Based on prior publications (Dalmat et al., 2019) and expert discussion to synthesize use cases based on analysis requirements, we defined eight main use cases for *Plasmodium* genetic data informing malaria control and elimination. For each use case, we identified specific functionalities needed for analysis. For a subset of use cases, we then outlined how these functionalities could be assembled into analysis workflows to obtain required results from the initial data.

*1. Identifying the molecular mechanism/origin of drug and diagnostic resistance*: Studies are initially needed to link parasite genetic variation to phenotypes of drug and diagnostic resistance, for example by sequencing parasites with corresponding data on *in vitro* response to antimalarials or with evidence of clinical failure of drugs or diagnostic tests. Related studies can identify potential polymorphisms of interest via signals of selection and identify how they are evolving and spreading at various temporal and spatial scales.
*2. Monitoring the prevalence/frequency of drug or diagnostic resistance markers:* Once genetic polymorphisms have been associated with resistance phenotypes, they can be used as markers of resistance. Marker prevalence (fraction of infections that contain parasites with the resistance marker) and/or frequency (relative abundance of the resistance marker in the parasite population) can be monitored for surveillance of resistance. Various statistical methods may be needed to estimate prevalence/frequency because polyclonal infections complicate otherwise simple analyses.
*3. Measuring human immune selection on the parasite population:* Identifying areas of the *Plasmodium* genome under immune selection can help identify potential targets for vaccines and related immunologic interventions such as monoclonal antibodies. Related to this, monitoring sequences of such targets can be useful in the context of large-scale implementation to evaluate for selection and monitor for potential escape mutations.
*4. Classifying outcomes in therapeutic efficacy studies (TESs) as reinfection, recrudescence or, in the case of P. vivax*, *relapse:* A TES performed by prospective evaluation of patients’ responses to treatment for uncomplicated malaria is the gold standard for assessing the efficacy of antimalarial drugs. Genotyping of participants with infections during follow up is needed to distinguish whether they failed therapy (recrudescence), were infected again (reinfection), or had a relapse of a dormant parasite.
*5. Estimating transmission intensity:* Population level measures derived from parasite genetic data, like other indicators such as parasite prevalence, may be used to estimate malaria transmission intensity for surveillance purposes such as stratification of interventions or to evaluate the effect of interventions.
*6. Estimating the connectivity and movement of parasites between geographically distinct populations:* Identifying whether and to what extent parasites from one geographic area are moving to another can help predict the spread of malaria, as well as the spread of parasites with specific concerning phenotypes such as drug resistance. This in turn can inform planning of the scale and timing of interventions. For example, it may be easier to reduce the burden of malaria in an isolated area compared to one that is more connected to areas of higher transmission. This information may be identifiable by comparing genetic measures between populations.
*7. Classifying malaria cases as locally acquired or imported from another population:* In elimination or pre-elimination settings, understanding if cases are imported is relevant to 1) assessing the feasibility of elimination (i.e. elimination is less feasible in areas with a high number of imported infections that may lead to local transmission) and 2) determining if local elimination has been achieved despite reported cases.
*8. Reconstructing granular patterns of transmission:* In areas of very low transmission, characterizing granular details of transmission events — such as chains of transmission between individuals/households, demographic groups at high risk of being infected with or transmitting malaria, hidden reservoirs of residual transmission, contribution of imported cases to local transmission, and the length of sustained transmission chains may help guide and evaluate targeted elimination strategies. This information may be identifiable through studies that sample infected people comprehensively and utilize high resolution genetic data to evaluate relationships between infections.

Each of these eight use cases requires several analysis functionalities to obtain the desired results from Plasmodium genetic and contextual data. We identified 14 distinct analysis functionalities that are required or useful for the eight use cases, with many functionalities shared by several use cases (Table 1). For example, estimating complexity of infection (COI, the number of genetically distinct clones within an infection) was identified as required or useful for all eight use cases. This makes intuitive sense, since analysis of genetic data from polyclonal infections often needs to be treated differently than that from monoclonal infections. Similarly, estimating phase (assigning alleles to particular clones within a polyclonal sample), population allele frequencies, and within-host relatedness were identified as useful for the majority of use cases. Thus, having a curated set of validated software tools to carry out these functionalities would support a diverse range of analyses. Other functionalities such as estimating copy number variation or multi-locus genotype frequencies were only needed for specific use cases.

**Table 1:** Mapping use cases to core functionalities.

| Use case | Analysis functionality |  |  |  |  |  |  |  |  |  |  |  |  |  |
| --- | --- | --- | --- | --- | --- | --- | --- | --- | --- | --- | --- | --- | --- | --- |
|  | Estimate COI | Phase | Estimate allele prevalence | Estimate allele frequency | Estimate multi-locus genotype prevalence | Estimate multi-locus genotype frequency | Estimate between-host relatedness / genetic distance (e.g. IBD, IBS) | Identify signs of selection | Estimate within-host relatedness / genetic distance (e.g. IBD, IBS) | Infer transmission networks | Infer geographic assignment | Calculate simple genetic metrics | Infer structure / clustering | Estimate copy number variation / deletions |
| Identifying the molecular mechanism/origin of drug and diagnostic resistance | X | X | X |  | X |  |  | X | X |  |  |  |  |  |
| Monitoring the prevalence/frequency of drug or diagnostic resistance markers | X | X | X | X | X | X |  |  |  |  |  |  |  | X |
| Measuring human immune selection on the parasite population | X | X | X | X |  |  |  | X |  |  |  |  |  |  |
| Classifying outcomes in therapeutic efficacy studies (TESs) as reinfection, recrudescence or, in the case of P. vivax, relapse | X | X |  | X |  |  | X |  | X |  |  |  |  |  |
| Estimating transmission intensity | X | X |  | X | X |  | X |  | X |  |  | X |  |  |
| Estimating the connectivity and movement of parasites between geographically distinct populations | X | X |  | X |  |  |  |  | X |  | X | X | X |  |
| Classifying malaria cases as locally acquired or imported from another population | X | X |  | X |  |  |  |  | X | X | X |  | X |  |
| Reconstructing granular patterns of transmission | X | X |  | X |  |  |  |  | X | X |  |  |  |  |

For three example use cases, we outlined how analysis workflows might be assembled from the component functionalities (Figure 2). During the process of outlining the workflows, several observations became noteworthy. First, obtaining seemingly straightforward results such as the frequencies of polymorphisms associated with drug resistance was more nuanced than generally appreciated. Second, there were often several viable paths to obtain a given result, based on features of the data (e.g. monoclonal vs. polyclonal population) and alternative analysis approaches. Third, within a given path, several software tools might interchangeably provide a given functionality, as discussed in the next section. Alternatively, some workflows are not sequential. They estimate key parameter values (e.g., COI, phase, and frequencies) using data on all samples jointly (i.e., they borrow information across samples to make up for relatively uninformative per-sample data) and would fail if polyclonal samples were separated from monoclonal ones. Finally, analysis approaches for some use cases (e.g. estimating transmission intensity) were less mature and although some analysis steps were clear, others remain poorly defined.

**Figure 2.**
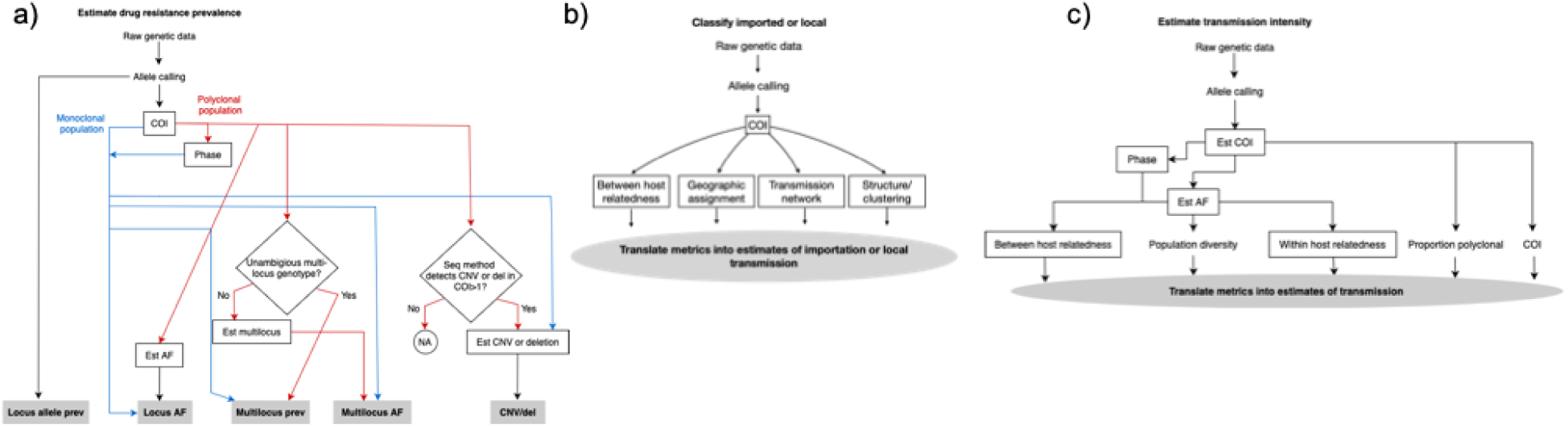
Analysis workflow for key use cases – (a) to estimate drug resistance prevalence, raw genetic data is processed through allele calling and subsequent steps vary depending on the complexity of infection (COI) and whether the population is monoclonal or polyclonal (including monoclonal and/or polyclonal infections). If the population is monoclonal, allele prevalence (prev) and allele frequencies (AF) at a single locus can be estimated directly and will be identical. Additionally, multi-locus prev, AF and copy number variation (CNV) or deletions (del) can be estimated directly if relevant. If the population is polyclonal and phasing is possible, the analysis follows the same pathway as for a monoclonal population. If phasing is not possible or not performed, additional steps are required depending on whether unambiguous multilocus genotypes can be obtained and whether the sequencing (seq) method utilized detects CNV or deletions in polyclonal infections; (b) to classify cases as imported or locally acquired raw genetic data is processed through allele calling, followed by estimation of COI. Then estimation of between host relatedness, geographic assignment, transmission networks and structure and/or clustering metrics can be translated into estimates of importation or local transmission. (c) to estimate transmission intensity, raw genetic data is processed through allele calling, COI and AF estimation. Subsequent metric estimation involves measuring the proportion of polyclonal infections in the population, between and within-host relatedness and population diversity, which can all be collectively translated into estimates of transmission. Alternative analysis pathways may be used depending on whether phasing is possible or performed.

### Landscaping of available analysis tools and mapping of tools to functionalities

To begin building a centralized resource for analysis tools to support the eight use cases, we curated and compared software characteristics of 40 tools (Supplementary Table 1). Twenty two of these tools were found to pass minimum inclusion criteria for evaluation (see Methods), and the characteristics of these tools are shown in Table 2, including implementation type, installation method, platform compatibility, and associated resources (e.g. website, publication or pre-print). It is important to note that this is not an exhaustive list; there are available tools that we did not evaluate.

**Table 2:** Tool landscaping matrix *(attached as .xlsx)*

Among these tools, the majority are implemented as R packages (14 tools) and others as standalone scripts implemented in R (2 tools), C/C++ or Python (3 tools), Perl (1 tool) and a Windows executable file (1 tool). Most tools are platform-independent, capable of running on multiple operating systems, but one tool is specifically designed for Windows. The installation methods for these tools vary widely: seven require manual installation from source files, six can be cloned and run locally from repositories, six are available via package managers like CRAN or BiocManager, and three require compilation. For some of the tools requiring compilers, efforts have been made to provide ‘R wrappers’ to decrease installation barriers (e.g. McCOILR, hmmibdr).

We identified the availability of the 14 analysis functionalities (defined in Table 1) in each tool (Table 3). We focused on tools that are actively being used as some of the tools were deemed obsolete or superseded, for example if installation was not possible during our evaluation, or if they have been superseded by newer tools that perform similar functionalities (these reasons are described in Table 2). The most common functionalities are estimating COI, available in eight different tools, estimation of SNP allele frequencies, which is supported by five tools, and estimation of between- and within-host genetic relatedness, which are available in four and two tools, respectively. Given the number of tools that perform similar functionalities, benchmarking them against standardized datasets will be an important next step to enable validation towards a curated list of best practices. There are also several gaps in current tool capabilities. For example, no existing tools are specifically designed to infer malaria transmission networks, and there is limited support for functionalities like phasing and geographic assignment covered by only one tool each.

**Table 3:** Tools to analysis functionalities.

| Tool | Analysis functionality |  |  |  |  |  |  |  |  |  |  |  |  |  |
| --- | --- | --- | --- | --- | --- | --- | --- | --- | --- | --- | --- | --- | --- | --- |
|  | Estimate COI | Phase | Estimate allele prevalence | Estimate allele frequency | Estimate multi-locus genotype prevalence | Estimate multi-locus genotype frequency | Estimate between-host relatedness / genetic distance (e.g. IBD, IBS) | Identify signs of selection | Estimate within-host relatedness / genetic distance (e.g. IBD, IBS) | Infer transmission networks | Infer geographic assignment | Calculate simple genetic metrics | Infer structure / clustering | Estimate copy number variation / deletions |
| FreqEstimationModel | X |  | X | X | X | X |  |  |  |  |  |  |  |  |
| MultiLociBiallelicModel | X |  | X |  | X |  |  |  |  |  |  |  |  |  |
| THEREALMcCOIL | X |  |  | X |  |  |  |  |  |  |  |  |  |  |
| coiaf | X |  |  |  |  |  |  |  |  |  |  |  |  |  |
| DEploidIBD | X | X |  |  |  |  |  |  | X |  |  |  |  |  |
| isoRelate |  |  |  |  |  |  |  | X |  |  |  |  |  |  |
| hmmIBD |  |  |  | X |  |  |  | X |  |  |  |  |  |  |
| dcifer |  |  |  |  |  |  |  | X |  |  |  |  |  |  |
| moire | X |  |  | X |  |  |  |  | X |  |  |  |  |  |
| MALECOT | X |  |  | X |  | X |  |  |  |  | X |  | X |  |
| DRpower |  |  | X |  |  |  |  |  |  |  |  |  |  | X |
| estMOI | X |  |  |  |  |  |  |  |  |  |  |  |  |  |

### Evaluating software standards

Software standards are a critical component in the development and maintenance of reliable, efficient, and interoperable software systems. These standards encompass various aspects of software engineering, including coding practices, documentation, testing, maintenance and accessibility. We categorized software standards into two broad groups: 1) those aimed at end-users, and 2) those aimed at developers (Table 4).

**Table 4:**
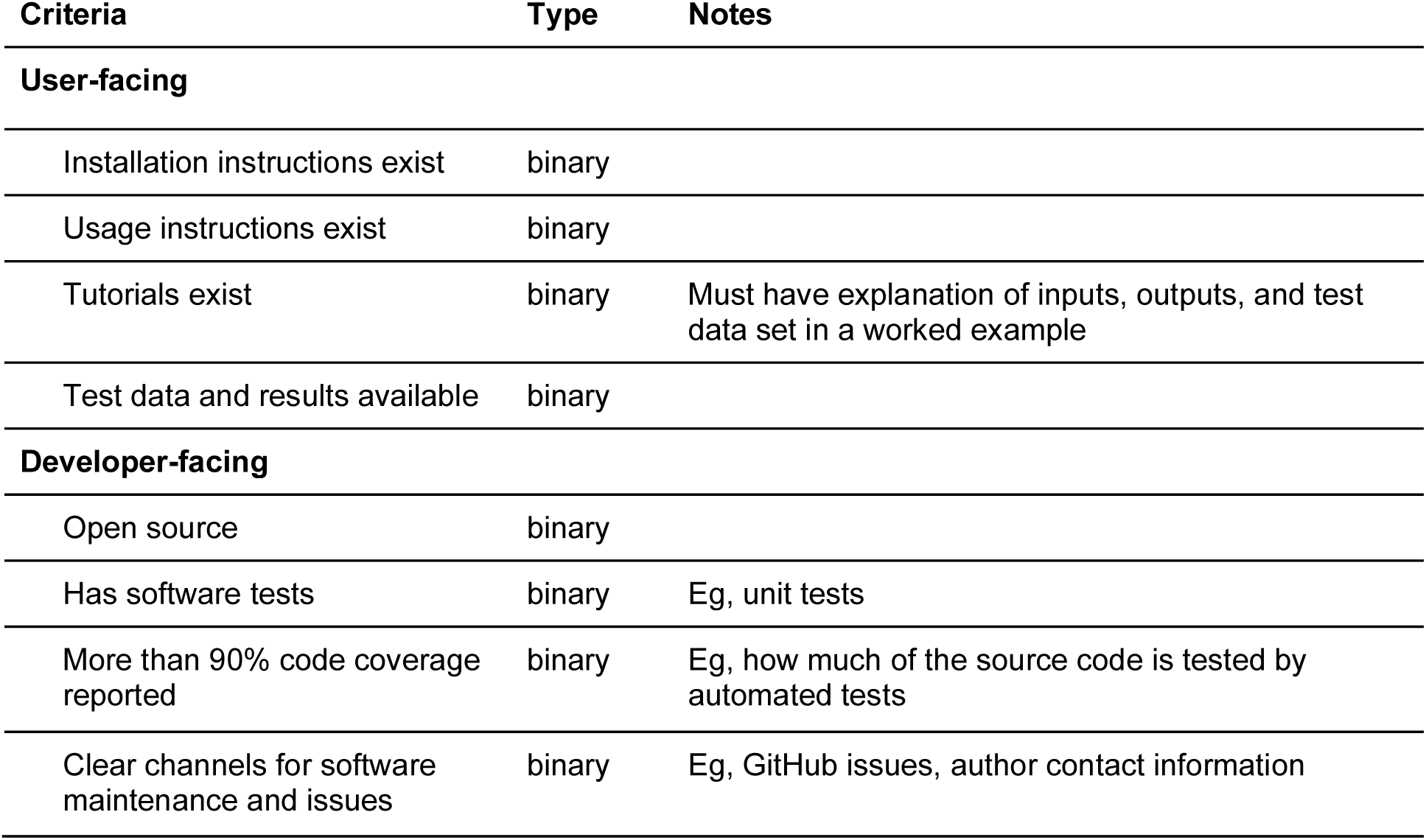
Software standards.

| Criteria | Type | Notes |
| --- | --- | --- |
| **User-facing** |  |  |
| Installation instructions exist | binary |  |
| Usage instructions exist | binary |  |
| Tutorials exist | binary | Must have explanation of inputs, outputs, and test data set in a worked example |
| Test data and results available | binary |  |
| **Developer-facing** |  |  |
| Open source | binary |  |
| Has software tests | binary | Eg, unit tests |
| More than 90% code coverage reported | binary | Eg, how much of the source code is tested by automated tests |
| Clear channels for software maintenance and issues | binary | Eg, GitHub issues, author contact information |

We applied all eight software standards to the 22 tools prioritized from the landscaping exercise. For each software standard, we used a scale from 0 to 2, where 0 indicates that the standard was not met, 1 indicates the standard was partially met (for example if usage instructions exist but are not comprehensive), and 2 indicates the standard was fully met. Overall, user-oriented software standards were generally met more consistently than developer-oriented standards (Figure 3).

**Figure 3.**
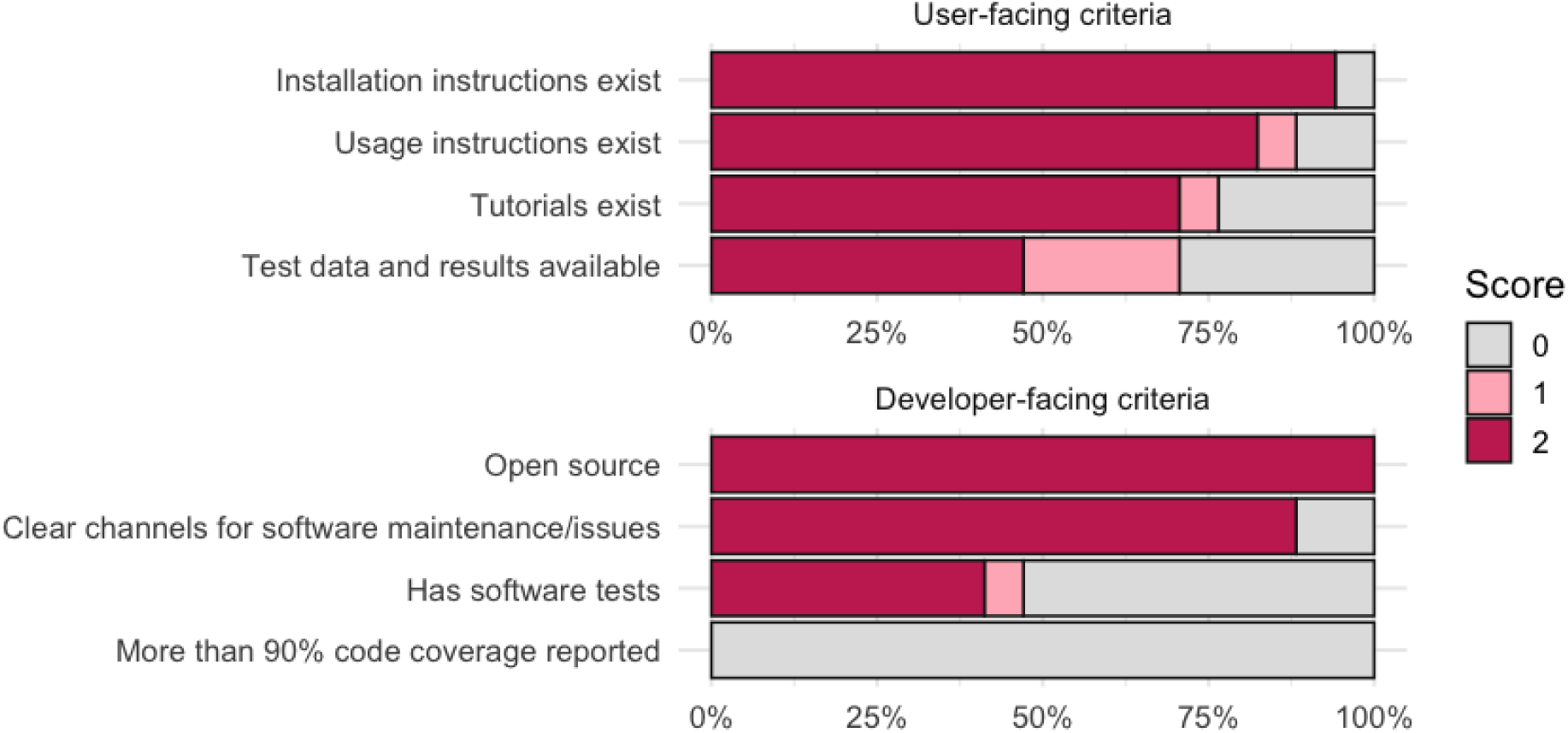
Evaluation of tools based on software standards for user- and developer-facing criteria.

Most tools provide basic installation instructions and usage guidance, whereas provision of tutorials and test data sets with expected results were not always available. For the developer-oriented software standards, although all tools were open-source and offered reasonably clear channels for software maintenance and issue resolution, very few provide adequate testing, and none achieve high code coverage. This points to a reasonable effort of making tools accessible and open-source for end-users, however there is still room for improvement in providing comprehensive usage resources and testing and documentation.

### Streamlining installation and managing system dependencies

Historically, a significant barrier to the adoption of software tools has been the challenges associated with their installation and the management of system dependencies. While tools may function correctly at the time of publication, differences in operating systems, software versions, and configurations often lead to incompatibilities that hinder successful installation or execution. Hidden system dependencies, in particular, can cause unexpected failures that are difficult to diagnose without advanced knowledge of the underlying software architecture. These obstacles can discourage end-users, especially those with limited technical expertise, reducing the accessibility and impact of the tools.

To address this, we developed the **PlasmoGenEpi r-universe** (<u>plasmogenepi.r-universe.dev</u>), a dynamic platform hosting R package repositories. This platform features an automated build process that incorporates platform-specific system dependencies, providing pre-compiled R packages to end-users and eliminating the need for local compilation. While this approach significantly improves usability and accessibility, further work on containerization would better facilitate fully reproducible analysis workflows that can operate seamlessly across diverse computational environments.

### Development of community resources

Community resources are essential for advancing malaria genomics by fostering collaboration, standardization, and accessibility in tool development and data analysis. To address this need, we developed PGEforge (<u>mrc-ide.github.io/PGEforge</u>) as a community-driven platform for end-users and developers of malaria genomic tools. PGEforge serves as a centralized hub for identifying and evaluating analysis tools, hosting example genomic datasets in widely used formats from both WGS and amplicon data and outlining analysis workflows tailored to key research questions. Additionally, it offers comprehensive and fully reproducible tutorials, initially developed for 10 analysis tools identified during RADISH23, that leverage the publicly available datasets hosted on the platform. This resource aims to streamline and enhance malaria genomics research through accessibility, standardization, and reproducibility.

## Discussion

The efforts described here represent an important step towards improving the usability of analysis tools for malaria genomic epidemiology, but substantially more work is needed to ensure useful results can be reliably obtained by a broad range of users. This work includes filling gaps in analysis functionalities, formally evaluating the accuracy of tools in estimating parameters, and creating a broader environment that facilitates flexible assembly of tools into accessible workflows. Mapping out workflows provides valuable insight into gaps currently underserved by existing software tools. Notable gaps include tools for *in silico* phasing of genomic data from polyclonal infections, which were identified as potentially useful across all eight defined use cases. Additionally, specialized tools are needed for specific use cases, such as the classification of outcomes in antimalarial therapeutic efficacy studies.

Where appropriate tools do exist, data on their accuracy are often limited (at best) to an initial evaluation by the original developers. Rigorous benchmarking of tools across different types of data, including assessment of performance in realistic situations that do not satisfy all model assumptions, will allow users and workflow developers to make informed choices on which tools are most appropriate for given applications. Standardized, curated datasets where the truth is known (e.g. from simulations or controlled experiments) along with well-defined performance measures will be an important step in facilitating routine benchmarking. For example, Guo and colleagues undertook comprehensive benchmarking of the accuracy of tools that estimate identity-by-descent using a simulated ground truth to show the impact of marker density on accurate IBD detection (Guo et al., 2024). Further benchmarking work is needed, including updated curation as the available set of tools expands and evolves.

Aspects important for end-to-end workflows, but not addressed during the RADISH23 hackathon, include evaluation of upstream steps of bioinformatic allele calling and downstream steps of communication and visualization of results. Choosing an appropriate bioinformatic pipeline to obtain allele calls is important since this will determine the type (e.g. SNPs, indels, and/or microhaplotypes) and quality of data used for subsequent analyses. Several pipelines exist for various types of sequencing data, e.g. built around GATK for obtaining SNPs and indels from whole genome sequencing data (Van der Auwera & O’Connor, 2020), or SeekDeep (Hathaway et al., 2018) or DADA2 (Callahan et al., 2016) for obtaining microhaplotypes from targeted sequencing data. However, there have been few efforts to perform systematic benchmarking of these pipelines. As with analysis tools, curated ground truth datasets will be useful in improving (e.g. optimizing tuning parameters), evaluating, and comparing pipelines. At the other end of the workflow, thoughtful consideration of how to best summarize and display results would substantially improve their accessibility and therefore public health impact. Providing several options for users, including summary tables and figures (e.g. https://genremekong.org/tools/grcmalaria-r-package-user-guide), potentially utilizing dynamic interfaces (e.g. (van Wyk et al., 2025), (Battle et al., 2024)), may be a worthwhile investment of resources. Linking all of these components together – allele calling pipelines, analysis tools, and display of results – is somewhat challenging at present. Creating a set of interoperability standards, e.g. agreed upon formats for inputs and outputs at various steps, would greatly facilitate assembly of these steps into complete workflows that start with raw genetic and contextual data and return results in interpretable formats. Similarly, creating example workflows e.g. well-commented scripts with associated documentation and tutorials that connect several disparate tools, can expand access to data analysis for specific use cases.

It will require some upfront effort to optimize, benchmark, and document the many steps required to guide raw sequence data through to meaningful results. However, such an investment is apt to pay large dividends, allowing harmonization of disparate components created via uncoordinated efforts. The end result would be a collaborative, transparent, and open platform of interoperable software, following the successful lead of this approach for other systems (e.g. QIIME, https://qiime2.org/). This model fosters community contributions, promotes healthy competition, and ensures that tools continue to evolve while remaining relevant and applicable. Important considerations to democratize access to such a platform will include documentation in several relevant languages beyond English and flexibility to install workflows locally or run them remotely on cloud-based platforms (e.g. (Data Sciences Platform, Broad Institute, 2024)).

In conclusion, a substantial body of work has been produced on analysis tools for *Plasmodium* genomic epidemiology. Thoughtful, targeted improvement of these existing tools in the context of an open ecosystem designed for organized growth has the potential to greatly extend their utility. We hope that the resources described here are a useful step in continued efforts towards this goal.

## Methods

### Landscaping of available analysis tools

We conducted a landscaping exercise of available tools that are commonly applied to *Plasmodium* genetic data. This included identifying, cataloging, and assessing tools. In order to limit the scope of this exercise during RADISH23, we applied the following constraints:

● Focus on downstream analysis tools. This includes tools whose primary goal is to extract signal from pre-processed data, but does not include tools that are primarily used within upstream bioinformatic steps, such as variant callers and quality filters.
● Focus on *Plasmodium* genetics, including both *P. falciparum* and *P. vivax*. We did not consider applications to mosquito genetics, despite some tools and techniques being applicable to both organisms.
● Focus on tools that are specifically engineered for *Plasmodium*. While there are many generic methods that can be applied to any type of genetic data (e.g. F-statistics and other population genetic metrics), we did not seek to evaluate these tools.

These constraints were necessary in order to make a meaningful contribution within a narrow timeframe, although we strongly advocate for future events expanding the scope of this exercise.

### Defining software standards for tool evaluation

When establishing objective criteria for evaluating software standards, we divided our approach into two categories: user-oriented standards and developer-oriented standards.

User-oriented software standards describe a set of best practices and guidelines that prioritize the needs and expectations of end-users. These standards cover interface design, usability, accessibility, and performance, which together create a cohesive and intuitive experience. Although central to commercial software development, user-oriented standards tend to be a blind-spot in academic software development where the incentive structure favors innovation over usability. We identified four standards as critical in promoting a positive user experience (Table 4), along with three characteristics that are desirable but not critical (Supplementary Table 2).

Developer-oriented software standards focus on ensuring that the software development process is efficient, maintainable, and scalable. This is important for the long-term viability of tools, the transparency of methods, and the potential to extend and modify existing frameworks as data and use cases change. We identified four standards as critical in promoting a robust development process (Table 4), along with four characteristics that are desirable but not critical (Supplementary Table 2).

### Compiling datasets of common genetic data input formats

We compiled empirical and simulated datasets of commonly used input formats that could be used as a reference set by the community. In addition, these same datasets were used across all tutorials developed during RADISH23, thereby ensuring results were reproducible and comparable between tools. We identified the following key features to ensure appropriateness and utility for downstream analysis tools:

- A subset of monoclonal and polyclonal samples, lab strains, and lab mixtures
- Various formats: VCF files, BAM files, tabular haplotypes in .csv
- A subset of polyclonal samples with within-host relatedness greater than 0
- Some amount of missing data

The data input formats derived were whole genome sequences, microhaplotypes, SNP barcodes, and *pfhrp2/3* gene deletion counts. All are freely available at the PGEforge GitHub repository at https://github.com/mrc-ide/PGEforge/tree/main/data.

### Whole genome sequencing data

We derived three VCF files from the MalariaGEN Pf3K project (https://www.malariagen.net/parasite/pf3k), selecting samples from Democratic Republic of Congo (DRC) (n=113), Vietnam (n=97), and laboratory strain mixtures (n=25). Each VCF contains 247,496 high-quality (VQSLOD>6) biallelic SNPs across all fourteen somatic chromosomes. The VCFs are sorted and accompanied by an index file. We provide associated contextual data for the samples, which includes Fws statistics from the MalariaGEN Pf7K dataset (https://www.malariagen.net/sites/default/files/Pf7_fws.txt) (except for the lab mixtures). These are deposited at https://github.com/mrc-ide/PGEforge/tree/main/data/wgs/pf3k.

We also simulated a dataset of samples with varying COI and within-host relatedness. Briefly, a simulated sample with a given COI, *K,* was created by randomly sampling *K* clonal haplotypes (Fws >0.95, (Manske et al., 2012)) from the Pf3K project (https://www.malariagen.net/parasite/pf3k), assigning these haplotypes to *j* ≤ *K* infectious bites, and if *j* < *K*, simulating meiosis with a constant recombination rate of 13.5kbp/cM (Miles et al., 2016). For each haplotype, proportions were randomly sampled from a symmetric Dirichlet distribution, and for each locus, read count data was simulated from a Beta-binomial distribution given the allelic proportions. Constant sequencing error was simulated in the allele counts at rates (REF to ALT, 10^-5^; ALT to REF, 5×10^−4^) broadly consistent with observation for the included sites. No variant calling error was simulated. In this way, we produced a VCF file with 40 simulated DRC samples, ranging from COI of one to four, with approximately half of the samples having within-host relatedness. Accompanying contextual data is provided as .csv and .bedfiles, which are deposited at https://github.com/mrc-ide/PGEforge/tree/main/data/wgs/simulated/pf3k-DRCongo-simA.

To derive BAM files, we subsetted the VCF calls to only *csp* (PF3D7_0304600), *celTOS* (PF3D7_1133400) and *ama1* (PF3D7_1216600) genes, and provide accompanying contextual data. These are deposited at https://github.com/mrc-ide/PGEforge/tree/main/data/wgs/labisolate_subset.

### Microhaplotype data

We provided microhaplotype data from 82 samples from Mozambique and lab strain mixtures from the study by (Tessema et al., 2020). The microhaplotype panel included 100 targets (91 diversity and 9 drug targets). The lab strain mixtures are from various combinations of seven *P. falciparum* lab strains, with some sequenced in replicate at 4 different parasite densities (10, 100, 1000, 10000 parasites per µl). The datasets and their accompanying contextual data can be found at https://github.com/mrc-ide/PGEforge/tree/main/data/amplicon.

We also simulated a Mozambique dataset to create 100 samples with 50 diverse targets sub-selected from the MAD^4HatTeR panel (Aranda-Díaz et al., 2024). Both PCR and Illumina error was simulated using in-house C++ scripts. Starting parasite densities were sampled from previous Mozambique data (Tessema et al., 2020), and PCR was simulated by taking the randomly sampled parasite density and simulating 23 rounds of PCR with a per-base error rate of 3.5e-06 for taq PCR rate and simulating chimeras. Illumina error was introduced by creating a Nextseq 3D7 reference data error profile. Haplotypes were randomly sampled from a previous Mozambique dataset, which was also used to generate a distribution of COI (da Silva et al., 2023).

### SNP barcodes

We derived SNP barcode data by subsetting the Pf3k WGS data from the samples from Vietnam and DRC described above to match the 101-SNP barcode panel from (Chang et al., 2019). The lab strain and lab mixtures dataset are subsetted to the 101-SNP barcode, with the mixtures created from various combinations of *P. falciparum* lab strains. We provide the accompanying contextual data with information on the genotypes that make up the mixtures and the COI.

We also simulated 101-SNP barcode data for 100 samples (50 from Bangladesh and 50 from Ghana) to include samples with a range of COI, as follows. A simulated sample was created by simulating superinfections by sampling the SNP barcode from each of these countries and selecting COIs based on the observed COIs in each country. Simulations were then created similarly to the details above for the Mozambique dataset but data was sampled from Pf7 dataset (MalariaGEN et al., 2023). We also provide an additional dataset where we removed indels. The datasets are provided at https://github.com/mrc-ide/PGEforge/tree/main/data/snp_barcode.

### pfhrp2/3 gene deletion counts

We provide *pfhrp2/3* gene deletion count data in tabular .csv format from the study by (Feleke et al., 2021) conducted in twelve sites in Ethiopia. The dataset can be found at https://github.com/mrc-ide/PGEforge/tree/main/data/pfhrp2-3_counts.

### Tutorial development

For each tool, we completed summary documents, installation aids and tutorials. Ten tools were completed during RADISH23, and guidelines were made available for future tutorial development by anyone in the community. These tutorials provide background information on each analysis tool, installation instructions and a fully worked-through analysis tutorial with code showing how an end user could use the tool to analyze data.

## Data availability

The empirical and simulated datasets produced during RADISH23 are freely available on the PGEforge GitHub repository at https://github.com/mrc-ide/PGEforge/tree/main/data. They were derived from publicly available data as described in <u>Methods</u>.

## Code availability

The resources developed during RADISH23 are freely available on the PGEforge website at https://mrc-ide.github.io/PGEforge/ and PGEforge GitHub repository at https://github.com/mrc-ide/PGEforge.

## Data Availability

The empirical and simulated datasets produced during RADISH23 are freely available on the PGEforge GitHub repository at https://github.com/mrc-ide/PGEforge/tree/main/data. They were derived from publicly available data as described in Methods. The resources developed during RADISH23 are freely available on the PGEforge website at https://mrc-ide.github.io/PGEforge/ and PGEforge GitHub repository at https://github.com/mrc-ide/PGEforge.

## Supplementary material

Supplementary Table 1: All tools landscaping matrix *(attached as .xlsx)*

**Supplementary Table 2:** “Nice to have” software standards.

| Criteria | Type | Notes |
| --- | --- | --- |
| **User-facing** |  |  |
| Informative error handling | binary |  |
| Multiple languages for tutorials | binary |  |
| Uses standard data input formats | binary | Eg decreases the need for the user to wrangle data into required input format |
| **Developer-facing** |  |  |
| Minimal dependencies | binary |  |
| Computationally efficient | binary |  |
| Modular code | binary | Eg split into functions |
| Well-annotated code | binary |  |

